# A systematic review of the performance of actigraphy in measuring sleep stages

**DOI:** 10.1101/2023.08.01.23293507

**Authors:** Hang Yuan, Elizabeth A. Hill, Simon D. Kyle, Aiden Doherty

## Abstract

The accuracy of actigraphy for sleep staging is assumed to be poor, but examination is limited. This systematic review aimed to assess the performance of actigraphy in sleep stage classification for adults. A systematic search was performed using MEDLINE, Web of Science, Google Scholar, and Embase databases. We identified eight studies that compared sleep architecture estimates between wrist-worn actigraphy and polysomnography. Large heterogeneity was found with respect to how sleep stages were grouped and the choice of metrics used to evaluate performance. Quantitative synthesis was not possible, so we performed a narrative synthesis of the literature. From the limited number of studies, we found that actigraphy-based sleep staging had some ability to classify different sleep stages compared with polysomnography.

## Introduction

Actigraphy has long been used to measure sleep-wake patterns in clinical and daily living environments. The broad adoption of actigraphy is driven by its relatively low cost, minimal user burden and ease of continuous recordings over weeks [1]. The American Academy of Sleep Medicine (AASM) published a meta-analysis on actigraphy for sleep detection in comparison with polysomnography (PSG) and concluded that actigraphy was suitable to assess total sleep time and sleep onset latency for clinical decision-making in adults with sleep disorders [2]. Extensive evaluations of actigraphy have been done on its population validity [3, 4], ecological validity [5] and device validity [6, 7] for sleep-wake classification.

Fundamentally, actigraphy and PSG measure different dimensions of sleep. PSG measures various physiological and movement signals, including brain activity (electroencephalography, EEG), ocular movement (electrooculography, EOG), muscle functions (electromyography, EMG) and heart rhythm (electrocardiography, ECG), with the combination of EEG, EOG, and EMG forming the basis of objective determination of sleep stages in line with published international guidelines [8]. In contrast, actigraphy measures peripheral limb movement alone, using activity as a proxy for wake and inactivity as a proxy for sleep, and by extension, has limited capacity to infer the stages of sleep due to the lack of physiological signals. Nonetheless, the use of actigraphy for sleep staging is relatively unknown. Understanding the performance limits of actigraphy-based sleep staging may open the door to investigate the association between sleep architecture and important clinical outcomes in large-scale biobank datasets, in which only actigraphy is available [9, 10].

The most relevant systematic review evaluated wearable sensing technologies, including actigraphy for sleep staging [11]. However, due to the broad coverage of the review, the depth of discussion related to each sensing modality was limited, and the review did not consider the technical parameters that could influence the algorithm performance for each device. Therefore, we set out to 1) systematically evaluate the validity of sleep staging using actigraphy and 2) analyse the effect of technical parameters on sleep staging performance.

## Method

This review was conducted according to the PRISMA (Preferred Reporting Items for Systematic Reviews and Meta-Analysis) guidelines [12]. The protocol was registered with the International Prospective Register of Systematic Reviews (PROSPERO) following the PRISMA-P guidelines (PROSPERO 2021 CRD42021237456) prior to commencement.

### Eligible studies

We included studies if they (1) compared the sleep staging agreement between actigraphy and PSG for adults, (2) reported agreement results on AASM sleep stages (wake (W)/non-rapid-eye-movement sleep (NREM) 1, 2, and 3 (N1, N2, and N3)/rapid-eye-movement sleep (R)) or performance on the composite of different sleep stages, and (3) scored the PSG manually or used an automated PSG scoring first and then corrected the scoring by a human rater. Studies were included regardless of the PSG recording montage used, provided the scoring followed the AASM guidelines.

Our exclusion criteria were: (1) editorials, reviews, or commentaries; (2) single-channel EEG as the reference when scoring sleep stages; (3) studies that only reported the agreement performance for sleep and wakefulness; (4) scoring using Rechtschaffen and Kales rules [13]. Supplement Table 1 lists the criteria for selecting eligible studies in detail.

### Search strategies

The searches were performed using MEDLINE (Ovid), Web of Science, Google Scholar and Embase (Ovid) on 25 July 2023. We also included preprints that appeared in Google Scholar as mitigate publication bias. The exact search terms for each database are specified in Supplement Section 3. Our search strategy was adapted from previous work [4], which aimed to assess the agreement between actigraphy and PSG using parameters related to sleep and wake patterns. To focus more on the performance of sleep staging, we made two changes to the search terms: (1) included more research-grade and consumer-grade actigraphy to capture a broader set of studies using different wrist-worn accelerometers (2) added “sleep stage” to the search terms to make the search results relevant to sleep staging. In developing the search syntax for each database, we first determined the Ovid syntax for Embase and MEDLINE, then translated the search terms to other databases.

### Study selection and data extraction

Literature search results were uploaded to Covidence (Covidence, Melbourne, Australia), an Internet-based systematic review program that assists reviewers in streamlining the review process. Covidence automatically flagged any possible reference duplication. One reviewer (HY) manually verified the duplication removal. HY also conducted the literature search and abstract screening. A second reviewer (EAH) independently verified that all the included abstracts meet the selection criteria. Full-text screening and data extraction were performed by both HY and EAH independently in duplicate. Initial reconciliation was undertaken by HY and EAH, with a third author resolving any outstanding disagreement.

The key data fields for extraction included sleep staging outcomes, device specifications (device type, data modality and sampling frequencies), data acquisition environment, and population characteristics (sample size, age, sex, and existing comorbidities). Information related to the sleep staging algorithms, including algorithmic techniques and validation methods, was also extracted. When multiple algorithms were reported in a study, data were extracted only for the algorithm with the best performance. If different datasets were used for algorithm development and validation, then the performance of the validation dataset was extracted. Supplement Section 1 lists detailed data items. The data extraction form was piloted on a subset of studies during the screening process.

### Data evaluation

The risk of bias was evaluated independently by HY and EAH using a modified QUADAS-2 (Quality Assessment of Diagnostic Accuracy Studies tool)[14]. Although QUADAS-2 has mainly been designed for primary diagnostic tools, the four key domains in QUADAS-2 are pertinent to the focus of the review: 1) patient selection, 2) index test, 3) reference standard, and 4) flow and timing. The key domains reflect whether all the participants received the same reference standard and participant inclusions throughout the analyses.

Domains 2 and 3 also reflected concerns around algorithmic validity. The following irrelevant signalling questions were removed from the original QUADAS-2: q2 in patient selection, q1 and q2 in index test, q2 in reference standard and q1 in flow and timing. Two additional questions were included to better assess algorithmic bias: q1 in the index test and q2 in the reference standard. The risk of bias is judged as “low”, “high”, and “unclear.” The risk of bias is judged as “low” if answers to all signalling questions are “yes”; otherwise, the risk of bias is judged as “high.” When the information available does not permit answering the signalling questions, then the risk is judged as “unclear.” Supplement Table 4 entails the modified QUADAS-2 in full.

## Results

Our literature search identified 5,590 records from the databases, 2,120 of which were removed due to duplication by Covidence (Figure 1). Another 3,390 records were removed after the abstract screening. Among the remaining 80 studies for full-text screening, wrong model input (n=25), conference abstract only (n=14), and wrong sleep parameters (n=14) were the top three reasons for exclusion before data extraction. In total, eight studies met the inclusion criteria and were included in this review. Supplement Table 5 summarises the exact reasons for removal after the full-text screening.

**Figure 1:**
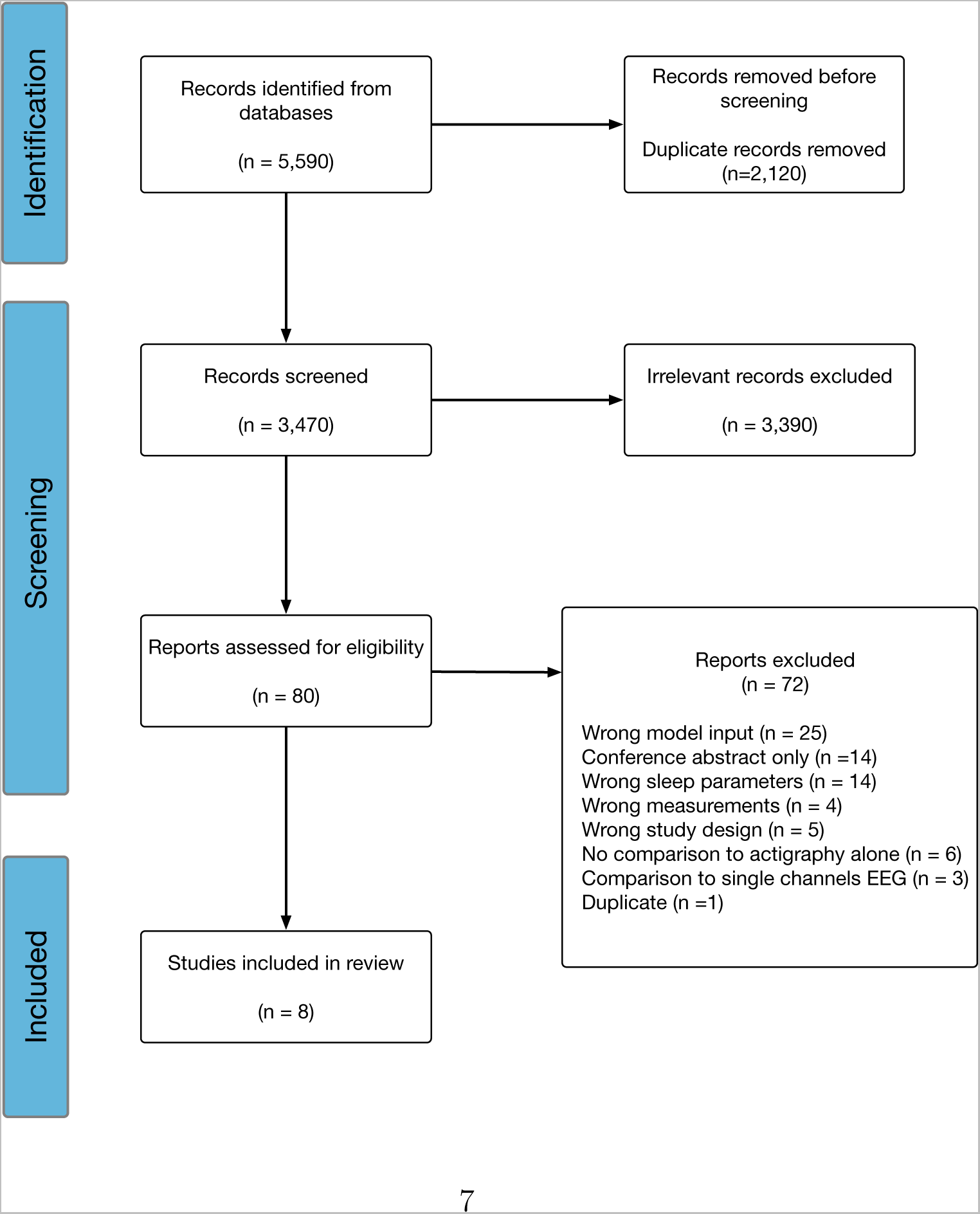
PRISMA flow diagram of studies screened and included to review the validity of actigraphy for the measurement of sleep stages.

### Study populations

The population characteristics are listed in Table 1. The majority of studies used data from healthy participants [15, 16, 17, 18, 19, 20, 21], whereas one study assessed the sleep staging performance in participants with sleep disorders [22]. The number of participants included in each study tended to be small. All but two studies included fewer than 100 participants in their assessment [17, 22], both of which reported the sleep classification performance using cross-fold validation without external validation. Even though Yuan et al. [21] also included more than 100 participants in their cross-fold cross-validation, the performance extracted is from external validation, which has only 52 participants.

**Table 1:**
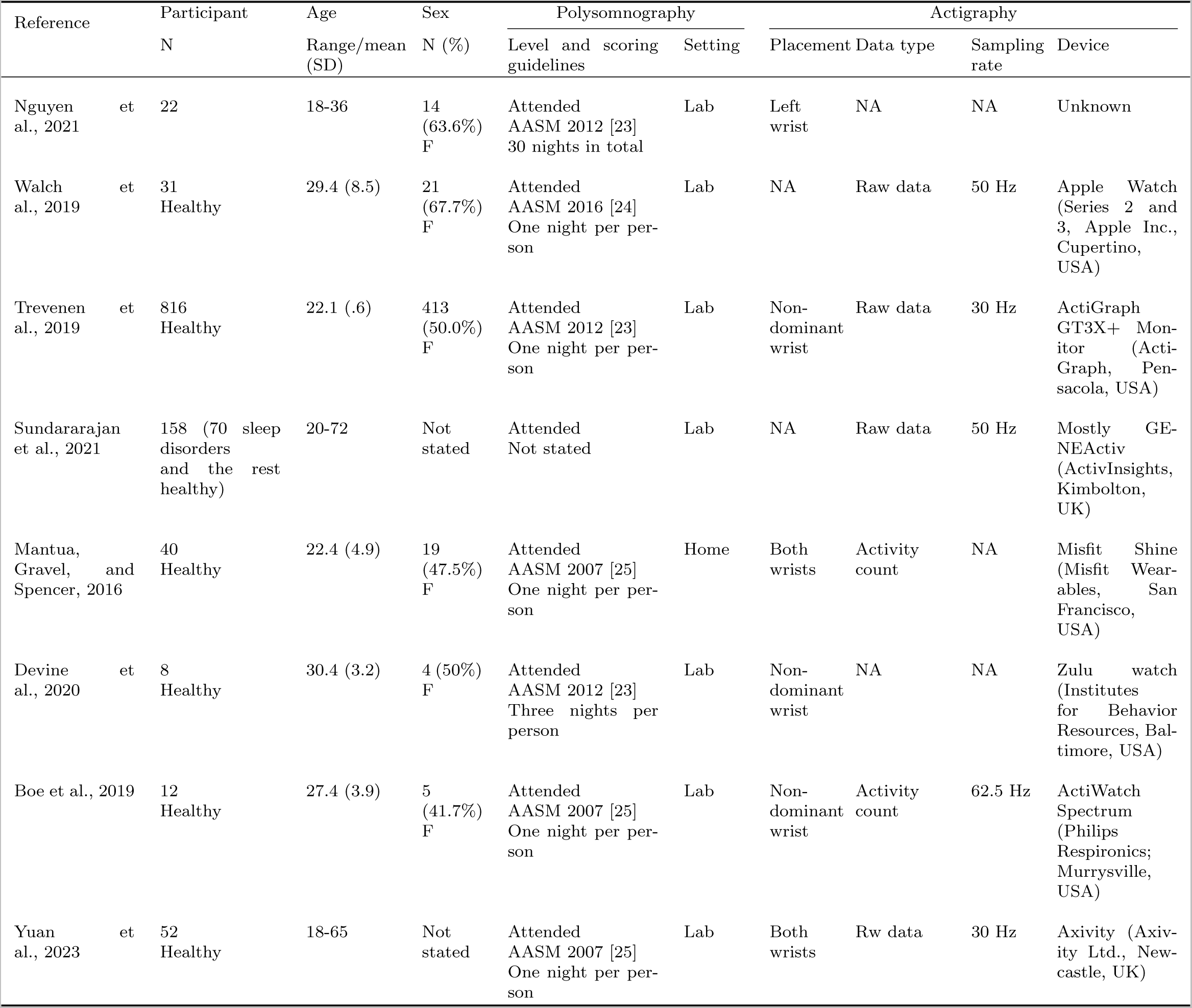
Study characteristics.

### Actigraphy devices

The full specifications of the actigraphy devices can be found in Table 1. Each study used a distinct type of device. Both research-grade and consumer-grade devices were used. There was no consensus on the device placement. Three studies placed the actigraph on the non-dominant wrist; one study placed the device on the left wrist; two studies placed the device on both wrists, and two studies did not report the device placement. Both raw data and activity count were used for sleep staging inference: four studies used the raw data, two studies used activity count, and two studies did not report the data type used. The actigraphy sampling rate for different devices ranged from 30 Hz to 62.5 Hz.

### Sleep staging algorithms

Table 2 shows the algorithms used and the corresponding performance from each study. Seven studies reported algorithm details. Studies that used commercial-grade devices relied on the proprietary algorithms from the vendors which do not reveal the process for sleep stage classification [18, 19]. Heuristic-based methods were not used by any study. Statistical learning methods were the predominant method class, including logistic regression [16], Hidden Markov Model [17], and tree-based methods [22, 20]. Deep learning techniques have also been used in some studies [22, 21].

**Table 2:** Sleep staging algorithms and performance. Wake: W; Non-rapid-eye-movement sleep 1, 2, and 3: NREM, N1, N2, and N3; rapid-eye-movement sleep: R; polysomnography: PSG.

| Reference | Algorithm | Validation | Sensitivity (%) | Specificity (%) | Accuracy (%) | F1 (%) | AUROC |
| --- | --- | --- | --- | --- | --- | --- | --- |
| Nguyen et al., 2021 | Not stated | Paired sample t-test | W: 27.9<br>N1+N2: 41.4<br>N3+R: 65.8 | W: 98.6<br>N1+N2: 62.1<br>N3+R: 51.6 | Average: 66.4 | NA | NA |
| Walch et al., 2019 | Logistic regression | Leave one out cross-validation | NA | NA | W: 60<br>NREM: 50.6<br>R: 33.2 | NA | NA |
| Trevenen et al., 2019 | Hidden Markov model | Leave one out cross-validation | NA | NA | NA | W: 61.0<br>N1: 10.0<br>N2: 52.2<br>N3: 63.8<br>R: 6.4<br>Average: 43.2 <sup>a</sup> | NA |
| Sundararajan et al., 2021 | Random forest | N-fold cross-validation | NA | NA | W: 60.7<br>N1: 1.0<br>N2: 57.9<br>N3: 27.9<br>R: 17.5 | W: 55.1<br>N1: 4.2<br>N2: 53.2<br>N3: 20.5<br>R: 12.3 | NA |
| Mantua, Gravel, and Spencer, 2016 | Proprietary algorithm | Not stated | Light: 23.4<br>Deep: 69.6 <sup>b</sup> | NA | NA | NA | NA |
| Devine et al., 2020 | Proprietary algorithm | Not stated | Light: 10.7<br>Deep: 84.2 | Light: 88.2<br>Deep: 30.0 | Light: 51.7<br>Deep: 48.7 <sup>c</sup> | NA | NA |
| Boe et al., 2019 | Bagged trees | Leave one out cross-validation | NA | NA | NA | NA | W: .82<br>N1&N2: .58<br>N3: .64<br>R: .47 <sup>d</sup> |
| Yuan et al., 2023 | Deep neural network | N-fold cross-validation & External validation | NA | NA | W: 54.3<br>N1: 24.6<br>N2: 38.4<br>N3: 54.7<br>R: 40.5<br>Average: 42.5 | NA | NA |

<sup>a</sup> The standard deviation for each class is: W (28.1), N1 (8.7), N2 (46.0), N (65.8), R (36.7).
<sup>b</sup> The average percentage errors are reported for the duration. Percentage errors: (Device duration - PSG duration)/PSG duration. Light: N1+N2; Deep sleep: N3+R.
<sup>c</sup> Each AASM PSG score is mapped from 0 (wake) to 3 (N3+R). Light sleep: any two-minutes bins with an average score between 1 and 2.55. Deep sleep: any two-minute bin with an average score greater than 2.5.
<sup>d</sup> The standard deviation for each class is: W (.18), N1&N2 (.05), N3 (.1), R (.09).

### Performance metric and assessment

Several performance metrics were used to report the sleep stage outcome, including sensitivity, specificity, accuracy, F1 and the area under the receiver operating characteristic (AUROC). Studies tended to report performance with different groupings of sleep stagings (Table 2). Hence, it is not possible to synthesise the evidence quantitatively. Only three studies reported the performance for all five AASM sleep stages [17, 22, 21]. The highest average five-class performance was 42.5% in accuracy [21], and 43.2% in F1 [17]. As the number of classes decreases, the reported performance increases. For instance, an accuracy of 66.4% was reported in a three-class setting (W vs N1+N2 vs N3+R) [15]. Studies using consumer-grade devices reported sleep staging performance by “light sleep” and “deep sleep” that were defined by the device vendors, making it impossible to compare sleep staging performance with other devices.

Method development studies used N-fold/leave-one-out cross-validation [16, 17, 22, 20, 21]. Only one method development study reported the performance on datasets that the algorithm has not seen [21]. However, device validation studies used statistic testing or directly calculated the performance metrics [15, 18, 19].

### Risk of bias of included studies

Overall, the risk of bias for the included studies based on the modified QUADAS-2 was limited (Table 3). The greatest risk of bias came from the participant selection, just one study with low risk, four studies with high risk and three studies with unclear risk. The next most common risk of bias was the index test having two studies with a high risk of bias, one with an unclear risk of bias and five with a low risk of bias.

**Table 3:** Risk of bias using assessment Quality Assessment of Diagnostic Accuracy Studies tool (QUADAS-2).

| Study | Risk of bias |  |  |  | Applicability concerns |  |  |
| --- | --- | --- | --- | --- | --- | --- | --- |
|  | Patient selection | Index test | Reference standard | Flow and timing | Patient selection | Index test | Reference standard |
| Nguyen et al., 2021 | (-) | ? | (-) | (-) | ? | (-) | (+) |
| Walch et al., 2019 | (-) | (+) | (+) | (+) | (+) | (+) | (+) |
| Trevenen et al., 2019 | ? | (+) | (+) | (+) | (+) | (+) | (+) |
| Sundararajan et al., 2021 | ? | (+) | (+) | ? | ? | (+) | (+) |
| Mantua, Gravel, and Spencer, 2016 | (-) | (-) | (+) | (+) | (+) | (+) | (+) |
| Devine et al., 2020 | (-) | (-) | (+) | (+) | (+) | (+) | (+) |
| Boe et al., 2019 | (+) | (+) | (+) | (+) | (+) | (+) | (+) |
| Yuan et al., 2023 | ? | (+) | (+) | ? | ? | (+) | (+) |
|  | Low Risk | (+) | High Risk | (-) | Unknown Risk | ? |  |

Several studies aimed to develop sleep staging algorithms and thus included limited information about participant inclusion and did not consider appropriate participant inclusion for clinical use. Two studies that carried a high risk of bias for the index test domain used consumer-grade devices [18, 19] that relied on proprietary staging algorithms, which could introduce bias. All but one study carried a low risk of bias for the reference standard PSG.

## Discussion

This review summarised the literature on the use of actigraphy in sleep staging. Eight studies were included, and only one study tested the validity of sleep staging in sleep patients. A large heterogeneity was found in the reporting of the performance in actigraphy-based sleep staging, especially around the sleep stage groupings and evaluation metrics, thus making it challenging to summarise existing evidence quantitatively. Nonetheless, based on the available studies, actigraphy-based sleep staging appears to have some ability to classify sleep stages.

Most of the evaluation of the actigraphy-based sleep staging has been conducted in young, healthy populations of limited size (*n <* 100). Only two of the included studies evaluated the sleep staging performance in more than 100 participants [17, 22]. Sundararajan et al. [22] was also the only study that evaluated the performance of actigraphy in patients with sleep disorders. Further evaluation of sleep staging performance in older adults and chronic conditions will help assess the clinical and scientific utility of actigraphy for sleep staging. However, the lack of open-access anonymous datasets requires researchers to collect their own datasets, which restricts the prospect of increasing the participant number and diversity for each study. Only one of the included studies used a dataset that was publicly available [22]. As a community, serious efforts should be put into making the data open access to allow for secondary data analysis.

The reporting of the sleep staging performance has been heterogeneous. Even though five sleep stages are defined by AASM, because of the difficulty in the sleep stage identification from actigraphy, several sleep stages are combined into one to improve the agreement with PSG. For example, Walch et al. [16] conducted the evaluation in a three-class setting (W/N1+N2/N3+R). While sleep stage grouping will help to obtain better performance, five-class staging performance needs to be reported to aid the comparison across different studies. Furthermore, studies included used a range of evaluation metrics for multi-class classification. Future research will benefit from adopting existing reporting guidelines to increase performance comparison across studies [26]. In particular, researchers should aim to report the staging performance at the individual level despite the grouping preference. In addition to the typical performance metrics like sensitivity and specificity, metrics robust to class imbalances, like F1 and Kappa scores, are encouraged.

Unlike sleep-wake classification for which heuristic-based methods are often used [27, 28, 29], all sleep staging methods reported here are based on statistical learning partly due to the difficulty of relying on pre-defined rules to infer sleep stages directly from wrist movement. Two studies leveraged recent advances in artificial intelligence (AI), such as deep neural networks, but, the model performance was limited [22, 21]. Data-driven methods like deep neural networks are particularly data-hungry and it would probably require thousands if not tens of thousands of nights worth of PSG data for deep neural networks-based methods to reach their potential.

Multi-modal methods for sleep staging predictions could help understand the value of each modality. Twenty-five references were excluded because of the wrong model input, and six studies were excluded because of no comparison to actigraphy alone. These groups of studies often used additional modalities such as heart rate or ECG to obtain better performance [30, 31, 32]. Reporting modality-specific performance in a multi-modal setting will inform the best modalities to use.

### Limitations

These results are likely to suffer from publication bias. It is possible that studies that could not classify sleep stages from actigraphy have not been published, particularly in individuals with chronic conditions for whom the classification will be more complex. Records were screened by a single reviewer in the initial abstract screening stage instead of two, as outlined in the study protocol, which may have inadvertently introduced some selection bias. However, all further steps in the review process were completed by two independent reviewers. Finally, this review did not consider the performance difference for studies with different validation methods due to the limited number of studies.

## Conclusion

Even though the included studies carry a limited risk of bias indicating the validity of sleep staging from actigraphy measurement, the existing performance remains low. Current evidence does not support or refute the validity of sleep staging from actigraphy due to the heterogeneity in the outcome reporting among the included studies.

### Practice points

• Actigraphy has some ability to di_erentiate between di_erent stages of sleep for young, healthy adults.
• Wide variability was found in the sleep staging grouping and evaluation metrics.
• Only half of the studies used evaluation metrics that were suitable for class imbalance.

### Research goals

• Adherence to rigorous reporting guidelines to aid performance comparison across studies: (1) reporting sleep stage-speci_c performance and the performance of desirable stage groupings and (2) reporting modality-specific performance and the performance of desirable multi-model setup.
• Subgroup analyses are needed to distinguish studies with di_erent research aims to assess sleep staging performance for the device, algorithmic, ecological and population validity.

## Data Availability

All data produced in the present work are contained in the manuscript

## Acknowledgments

HY, and AD are supported by Novo Nordisk. AD is additionally supported by Swiss Re, Wellcome Trust [223100/Z/21/Z], and the British Heart Foundation Centre of Research Excellence (grant number RE/18/3/34214).

SDK is supported by the NIHR Oxford Health Biomedical Research Centre, Health Technology Assessment Programme, Efficacy and Mechanisms Evaluation Progresso, and Programme Grant for Applied Research, and the Wellcome Trust. The views expressed are those of the authors and not necessarily those of the NHS, the NIHR or the Department of Health.

For the purpose of open access, the author has applied a CC-BY public copyright licence to any author accepted manuscript version arising from this submission.

## Data and materials sharing

Data extraction forms and references excluded after full-text screening can be found in the Appendix.

## Supplements

### 1. Data extraction item

- Authors, Reference number (text, number)
- Sample size (number)
- Funding sources (text)
- PSG type (text): at home recording or in a lab?
- PSG scoring rubric (text): R&K or AASM standard+year
- PSG channel/montage (text)
- PSG level if applicable: Level I/II (attended/unattended)
- Scoring method (sing-choice): manual, semi-automated, automated.
- Population type (text): age, population type (e.g. have sleep disorders or not),
- Recruitment approach (text)
- Device type/brand/mode
- Device placement
- Actigrpahy sampling rate
- Best performing algorithm name (text)
- Data source: activity count or raw accelerometery
- Sensitivity, specificity, accuracy and F1 for N1, N2, N3, combined NREM, R, and W.
- Balanced accuracy, kappa and F1 scores for all four sleep stages and W.

### 2. Eligibility criteria

**Table 1:** Eligibility criteria: accelerometer (ACC) and polysomnography (PSG).

|  | Inclusion | Exclusion |
| --- | --- | --- |
| Literature language | English | Not English |
| Literature type | Empirical studies | Non-empirical works such as:<br>• Editorials<br>• Reviews<br>• Commentaries |
| Study design | Studies with parallel ACC and PSG. | Studies that have only ACC or PSG. |
| Populations | Adults age $\geq 18$ years | Pediatric or adolescent populations with age $< 18$ years |
| Measurements | Wrist-worn ACC and (PSG or EEG). Level I/II (attended/unattended) PSG. | • Studies that do not have ACC<br>• Studies that only has either ACC or (PSG or EEG)<br>• Studies that have both ACC and only single-channel EEG. |
| PSG scoring | Only consider manual or semi-automated (initially scored automatically and then corrected by a human rater) | fully-automated scoring |
| Sleep parameters | Should at least report one of the following<br>• N1<br>• N2<br>• N3<br>• REM (R)<br>• NREM<br>• Other metrics, being composed by any of the above-mentioned measures, that are more specific than sleep. | Only reports sleep vs wake results or does not report any of the following<br>• N1<br>• N2<br>• N3<br>• REM (R)<br>• NREM<br>• Other metrics, being composed by any of the above-mentioned measures, that are more specific than sleep. |

### 3. Search terms

**Table 2:**
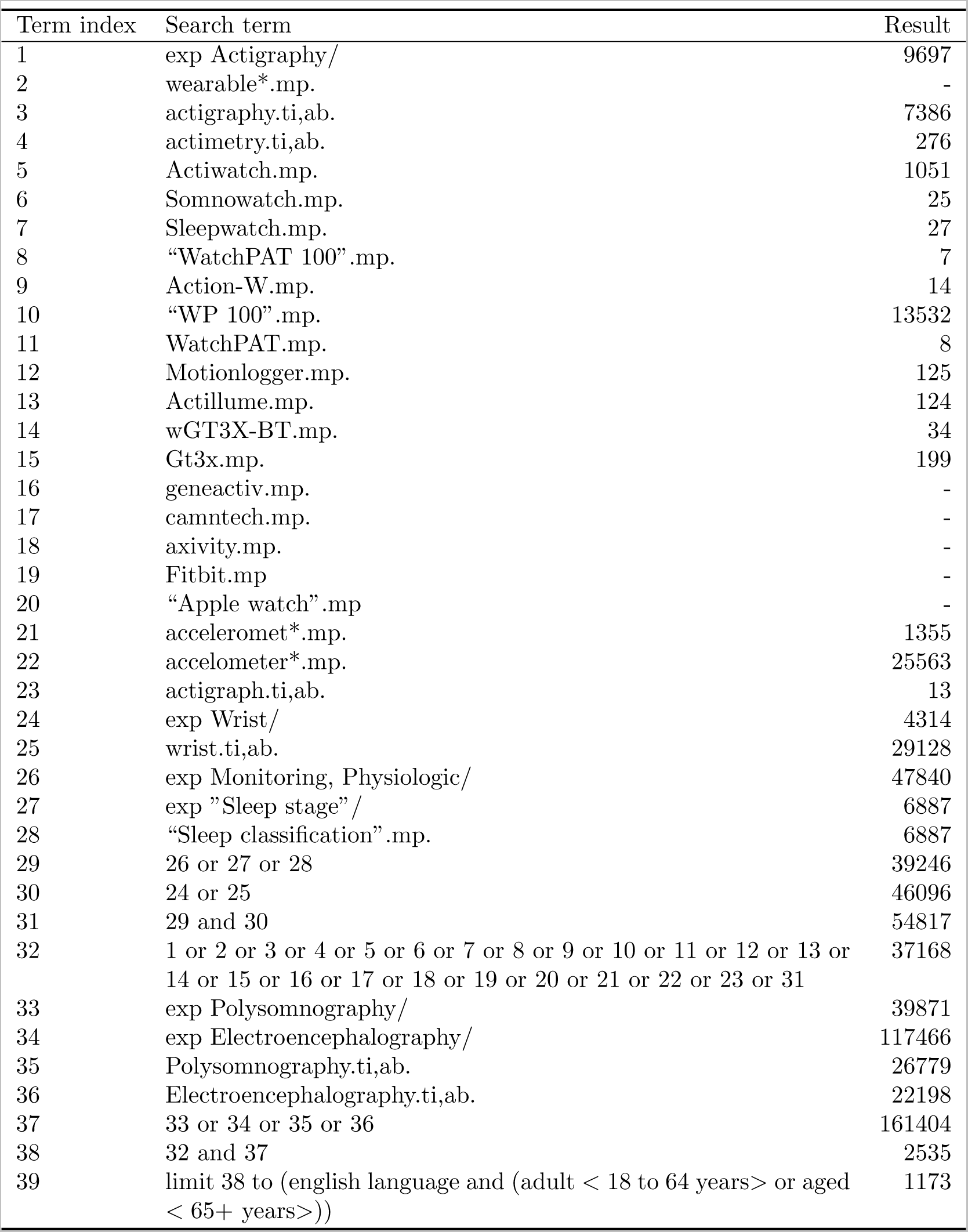
Search terms and results (Ovid).

**Table 3:**
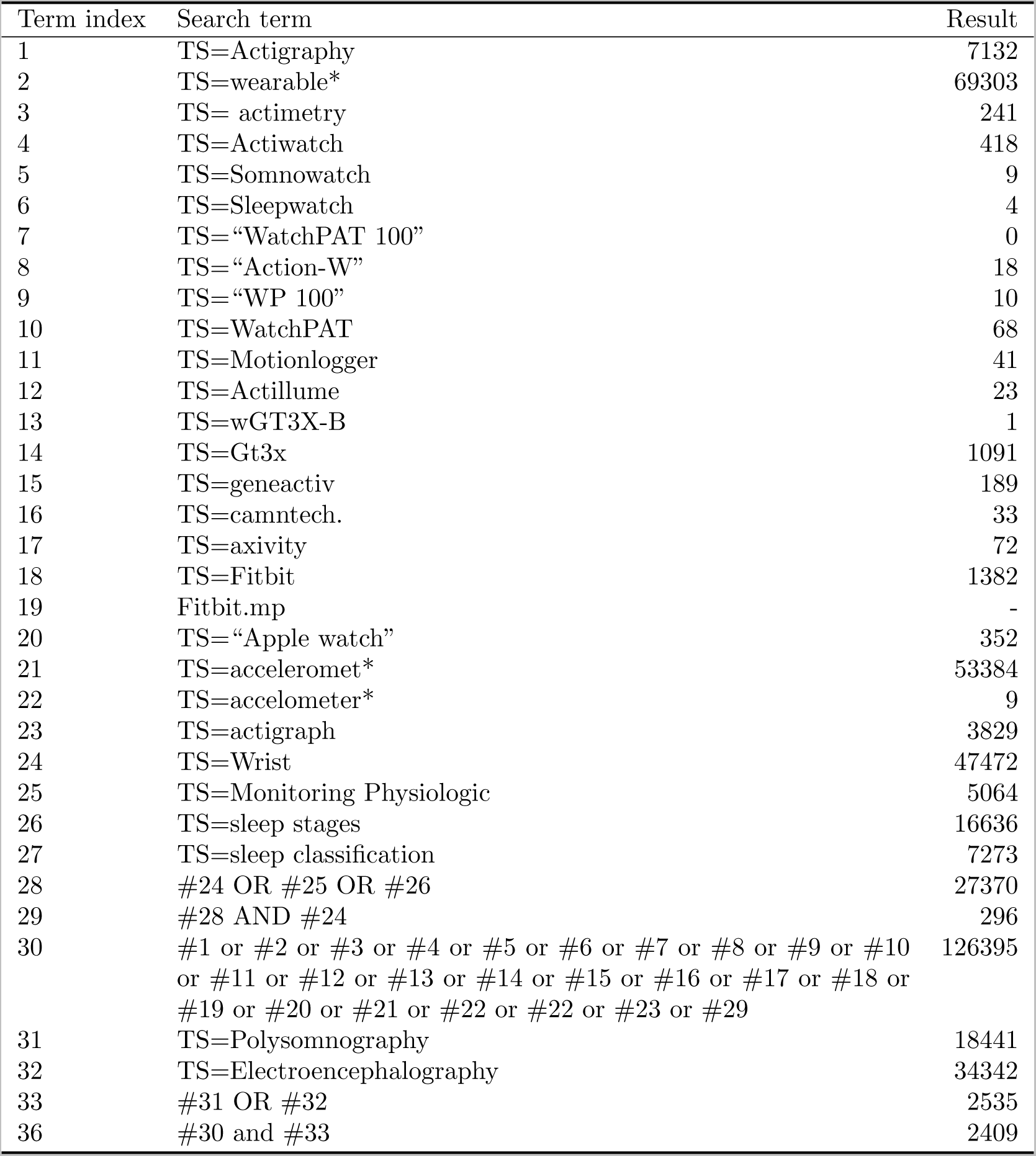
Search terms and results for Web of Science.

#### Embase and Medline

The above strategy is adopted from the strategy used in [1]. The search terms were used for both Embase and Medline. When performing the the search, the time limit for Embase was 1974 to present and the time limit for Medline was 1946 to present. The search terms and results are shown in Supplement Table 2

#### Web of Science

The search syntax was adopted from the Embase syntax. The search terms and results are shown in Supplement Table 3

#### Google Scholar

Stop at page 20 because no results are relevant on page 21. 10 links per page. (Actigraphy OR wearable OR actigraphy OR Actimetry OR Actiwatch OR Somnowatch OR Sleepwatch OR “WatchPAT 100” OR “Action-W” OR ‘WP 100” OR WatchPAT OR Motionlogger OR Actillume OR “wGT3X-BT” OR Gt3x OR geneactiv OR camntech OR axivity OR Fitbit OR Apple watch OR accelerometer OR accelerometry OR actigraph OR (Wrist AND ((Monitoring OR Physiologic) OR Sleep stage OR Sleep classification)) AND (Polysomnography OR Electroencephalography)

We put all the references into EndNote to search for abstracts before uploading them to Covidence for further processing.

**Table 4:** Modified QUADAS-2 form. Bold text questions are newly added.

| Domain | Patient selection | Index test | Reference stan-<br>dard | Flow and Timing |
| --- | --- | --- | --- | --- |
| Description | Describe methods of patient selection: Describe included patients (prior testing, presentation, intended use of index test and setting): | Describe the index test and how it was conducted and interpreted: | Describe the reference standard and how it was conducted and interpreted: | Describe any patients who did not receive the index test(s) and/or reference standard or who were excluded from the 2x2 table (refer to flow diagram): Describe the time interval and any interventions between index test(s) and reference standard: |
| Signalling questions (yes/no/unclear) | 1. Was a consecutive or random sample of patients enrolled? 2. Did the study avoid inappropriate exclusions? | **1. Was the index test obtained using a known algorithm?** | 1. Is the reference standard likely to correctly classify the target condition? **2. Was the reference standard interpreted using all participants as the test sample?** | 1. Did all patients receive a reference standard? 2. Did all patients receive the same reference standard? 3. Were all patients included in the analysis? |
| Risk of bias: High/low/unclear | Could the selection of patients have introduced bias? | Could the conduct or interpretation of the index test have introduced bias? | Could the reference standard, its conduct, or its interpretation have introduced bias? | Could the patient flow have introduced bias? |
| Concerns regarding applicability: High/low/unclear | Are there concerns that the included patients do not match the review question? | Are there concerns that the index test, its conduct, or interpretation differ from the review question? | Are there concerns that the target condition as defined by the reference standard does not match the review question? |  |

#### Excluded studies

**Table 5:**
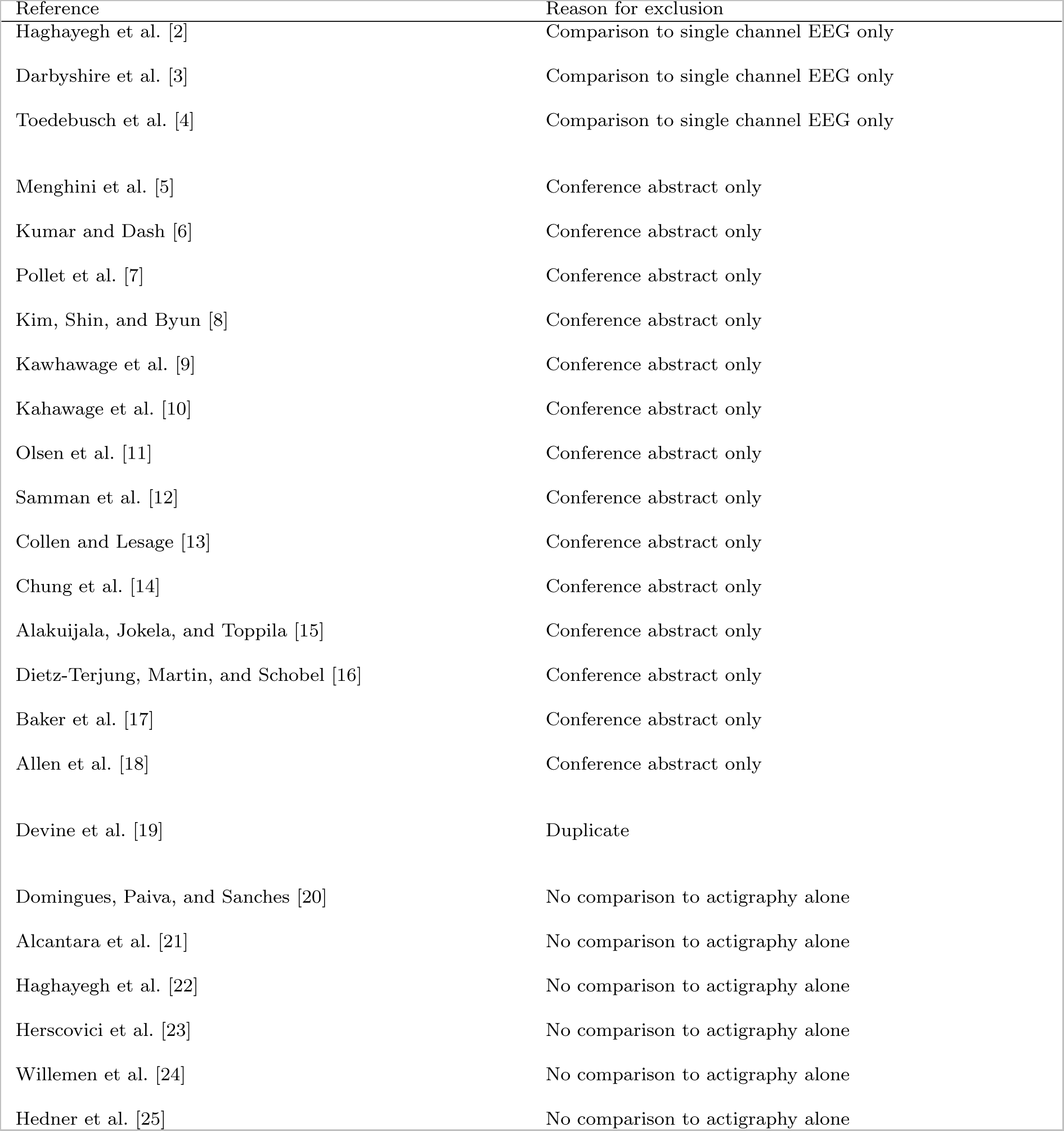
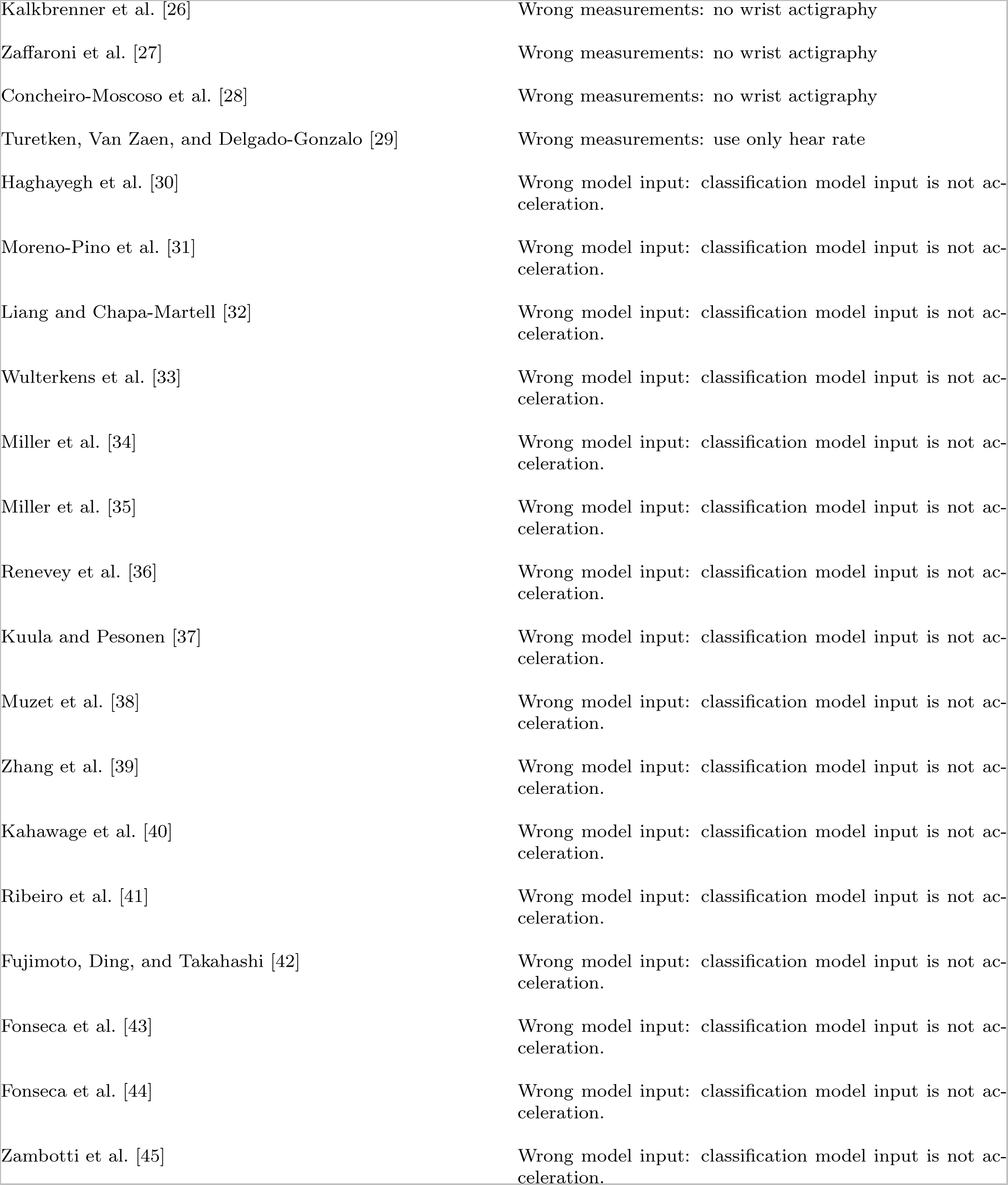
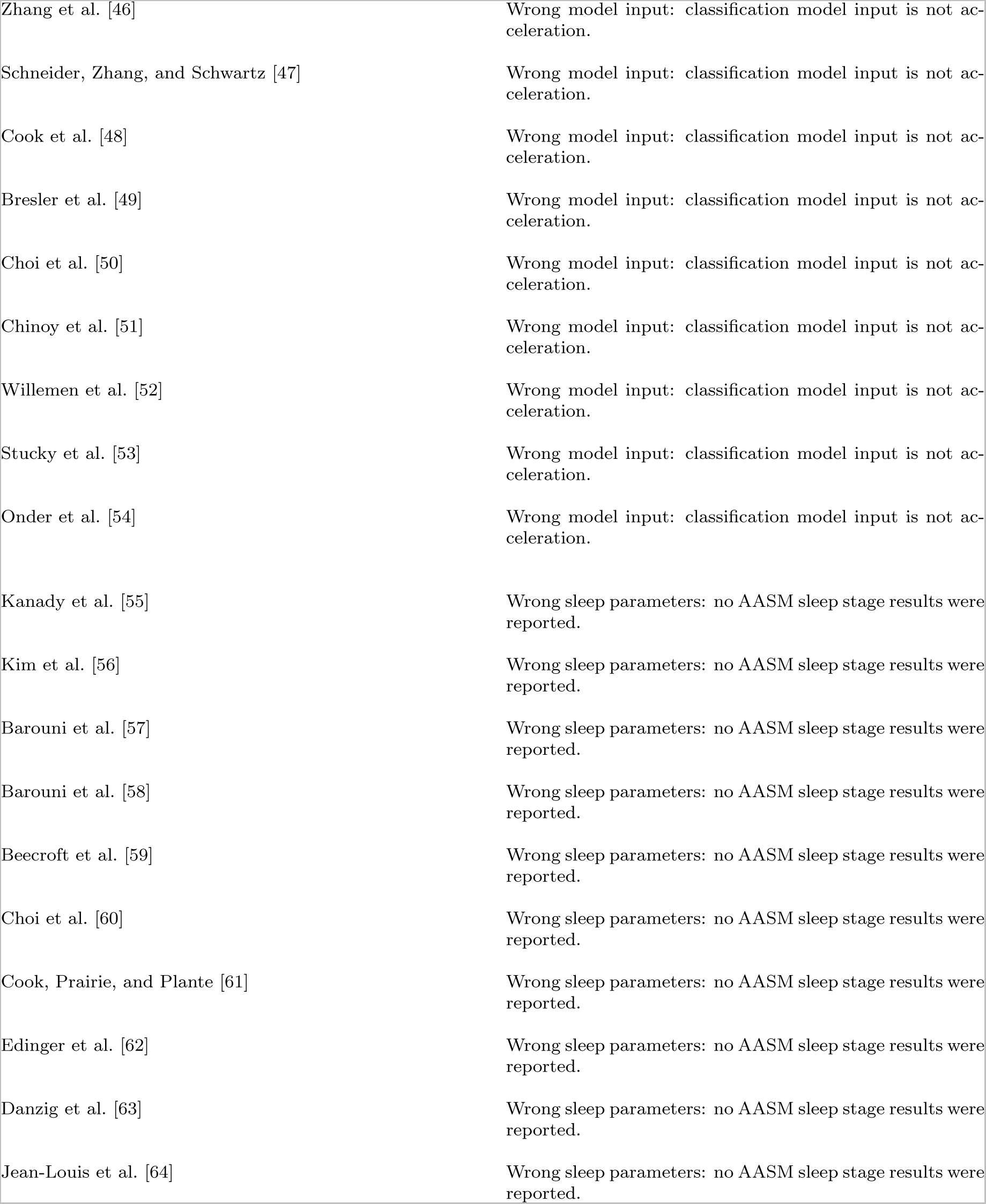
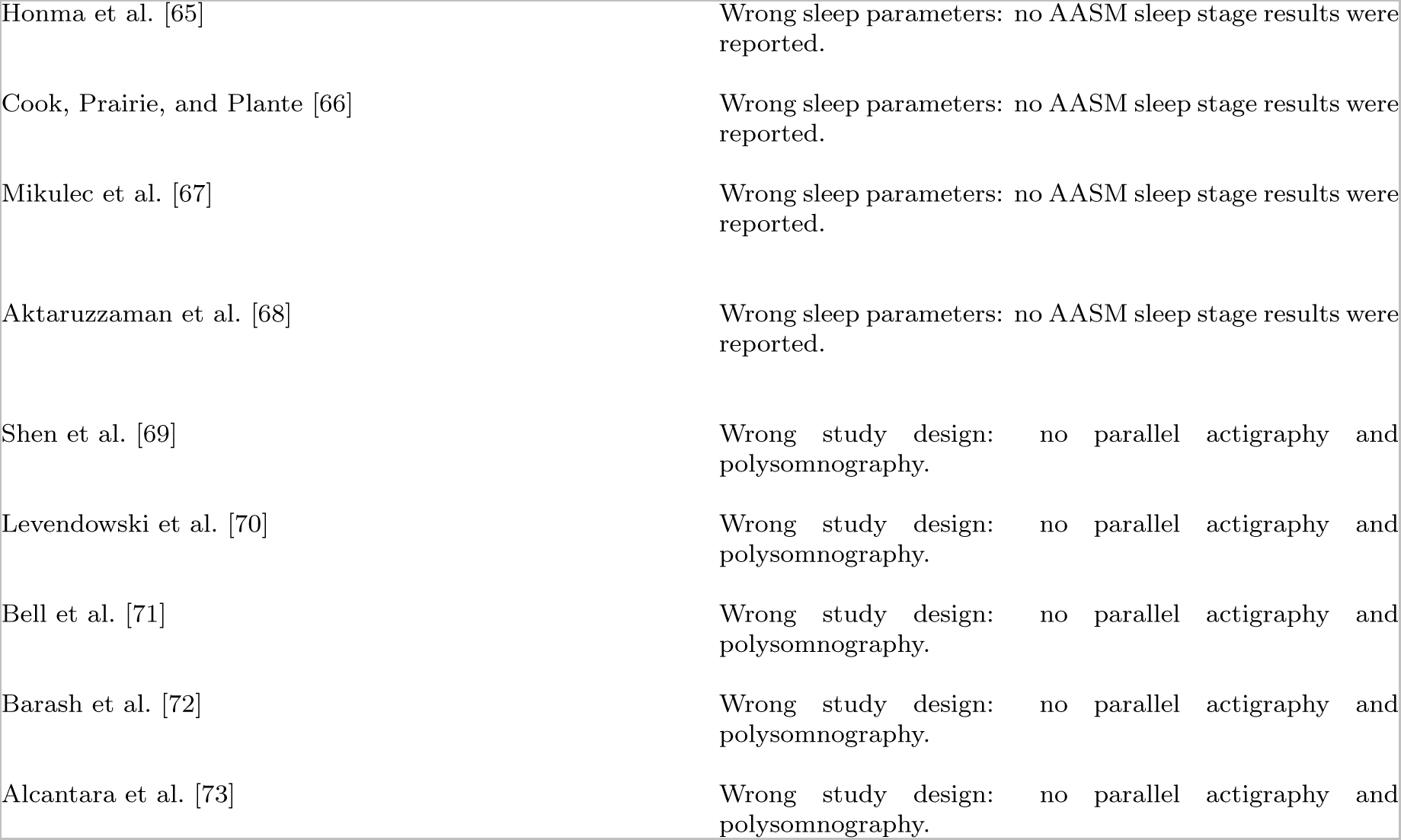
Exclusion reasons for studies that might be eligible. AASM: American Academy of Sleep Medicine.

## PRISMA 2020 Main Checklist

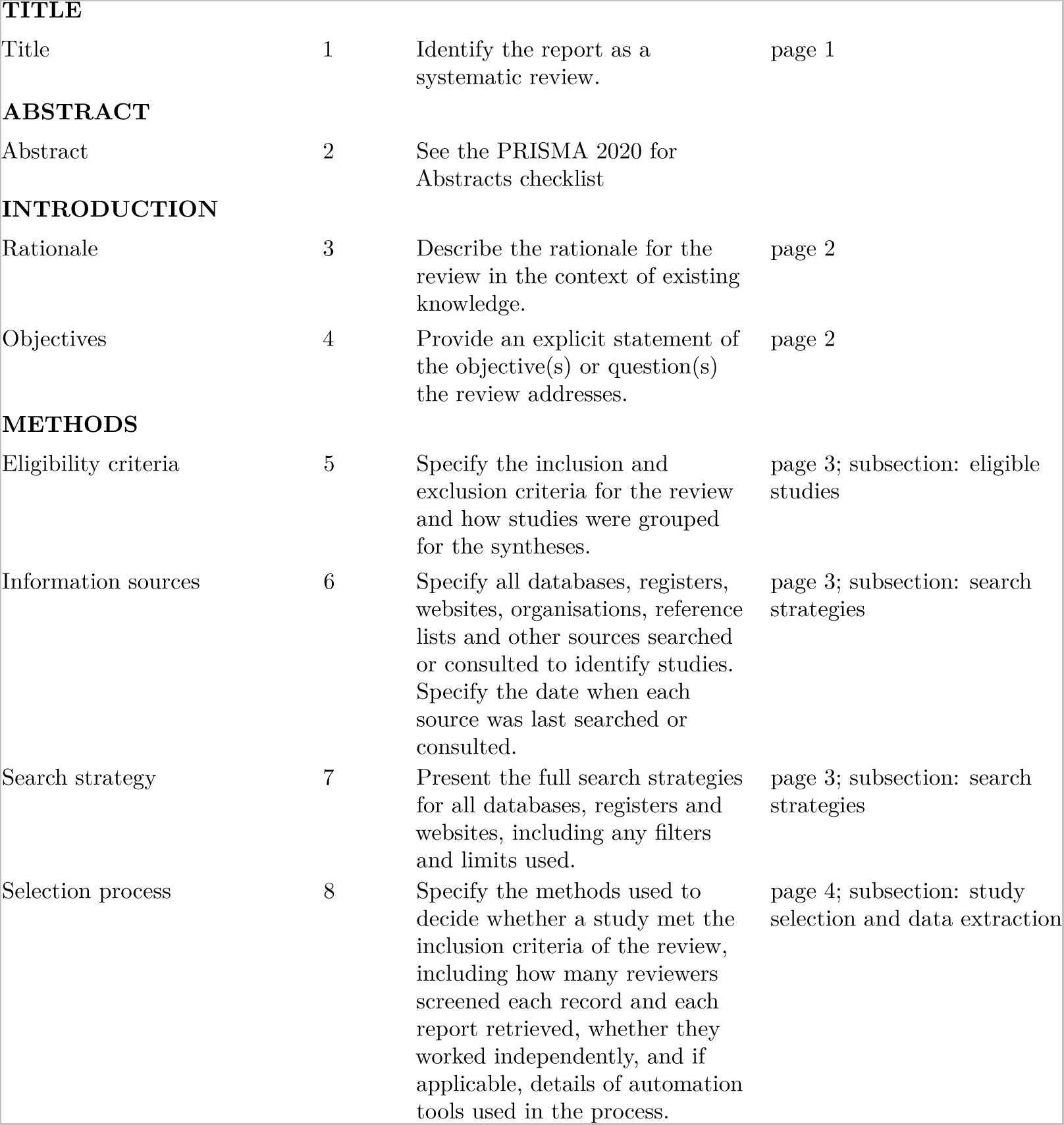
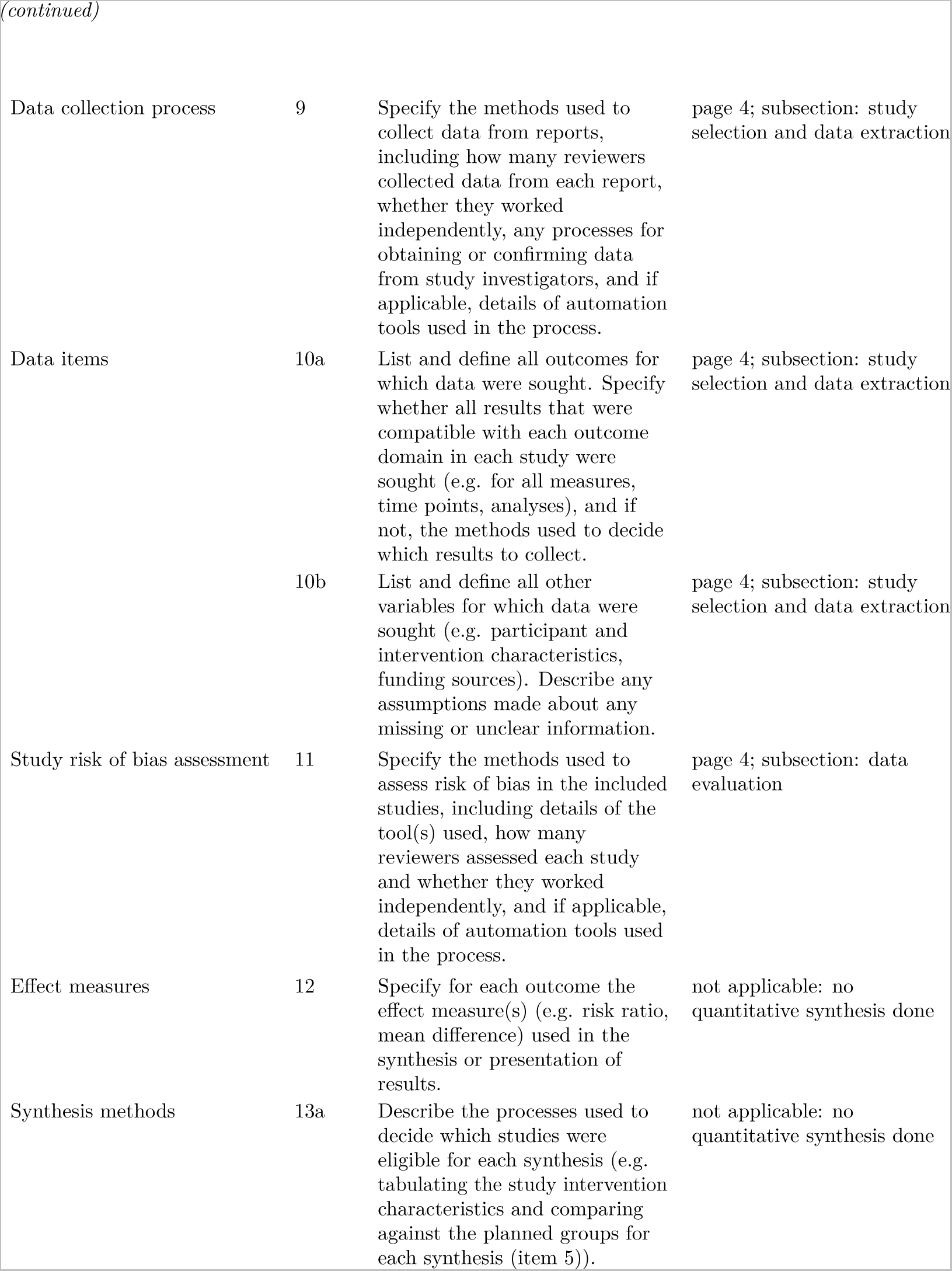
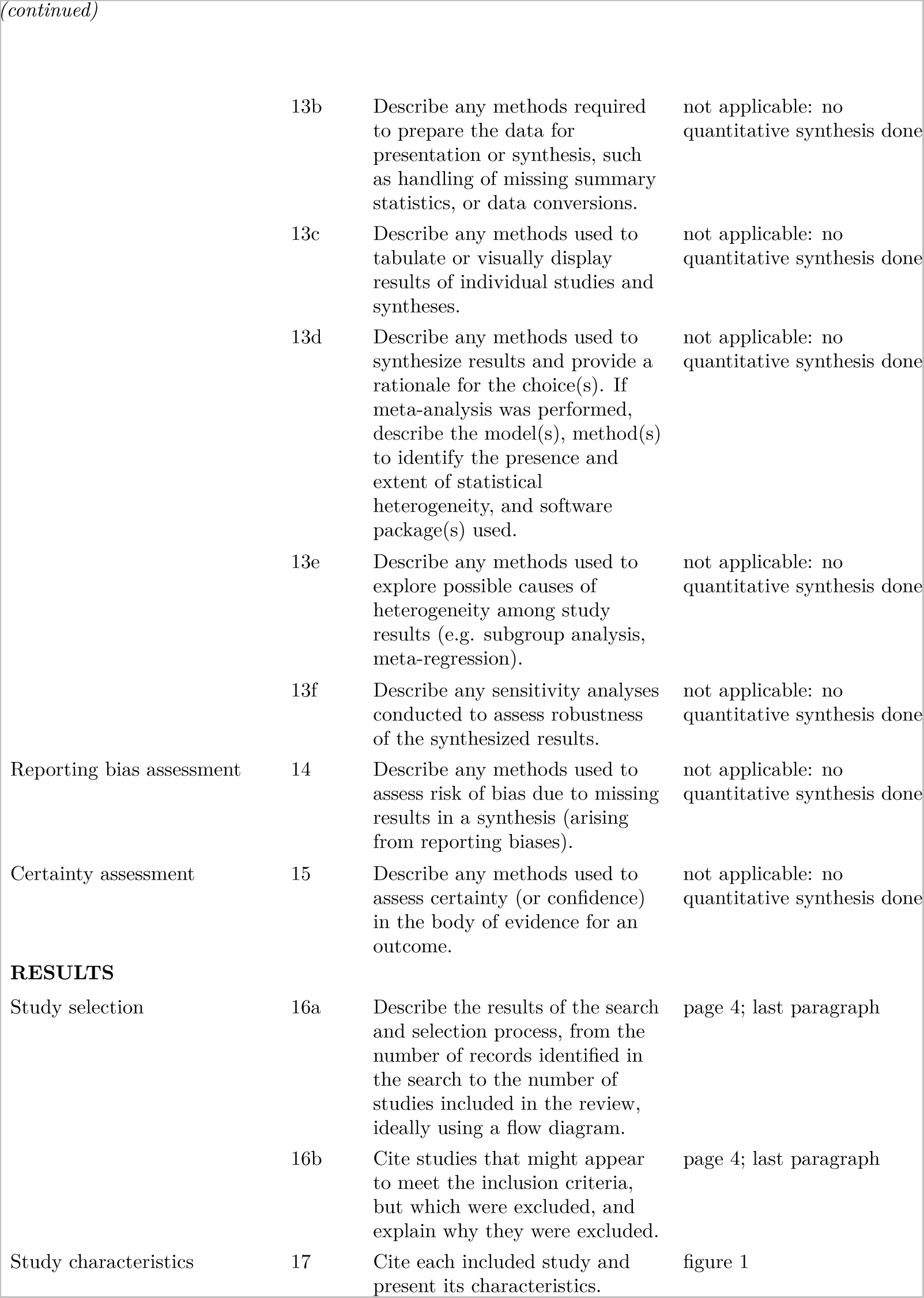
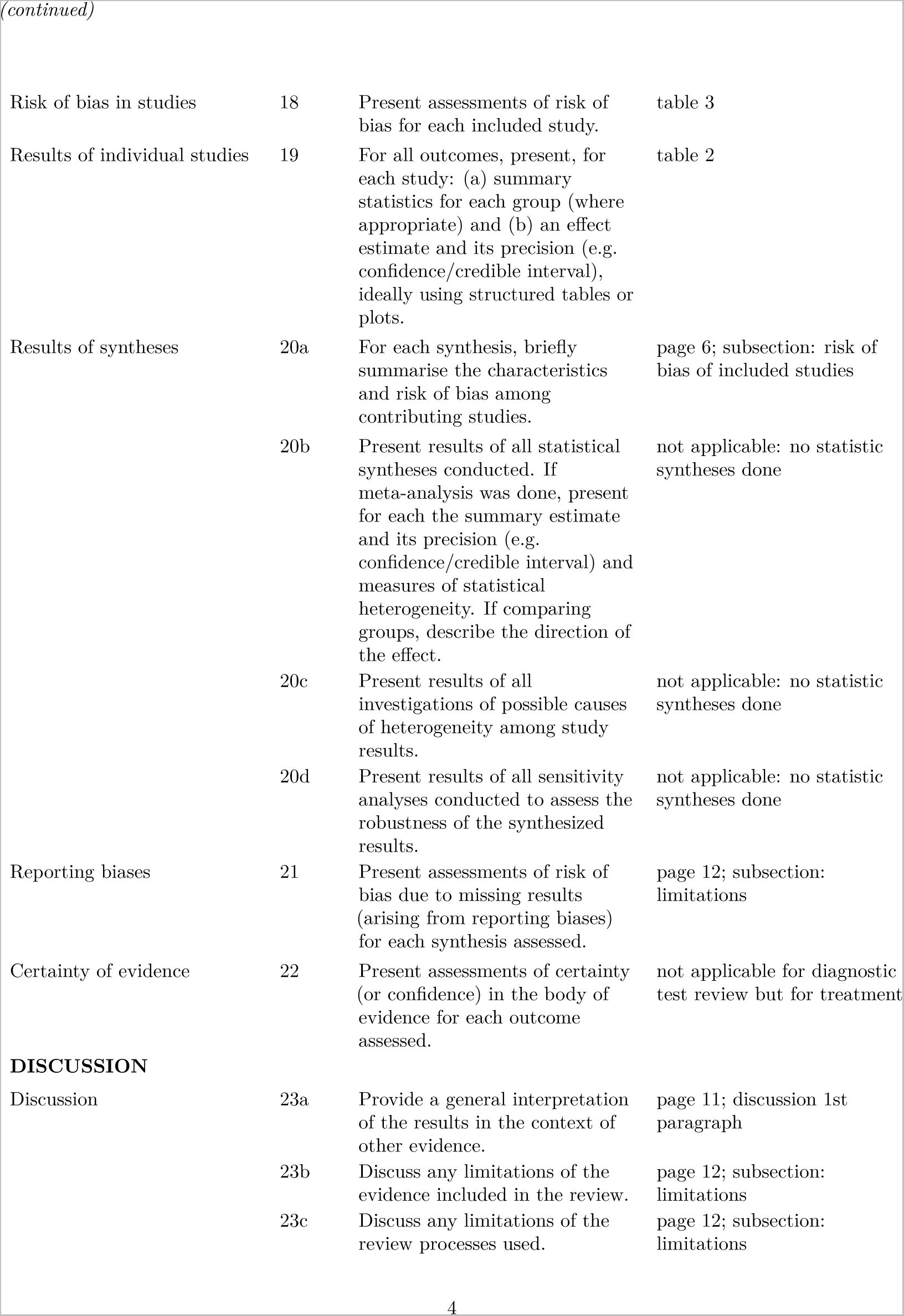
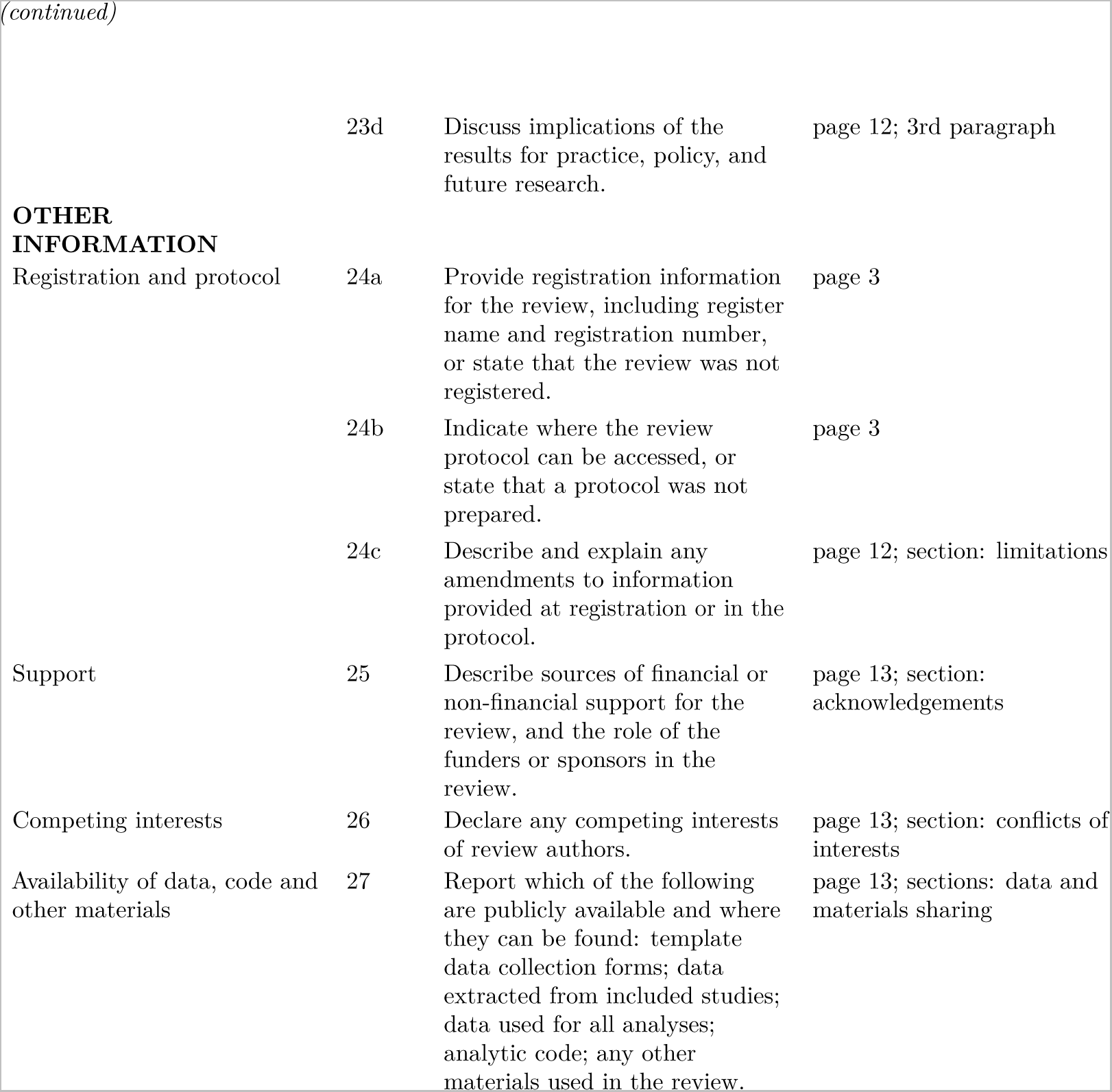

TITLE
|  |  |  |  |
| --- | --- | --- | --- |
| Title | 1 | Identify the report as a systematic review. | page 1 |

ABSTRACT
|  |  |  |
| --- | --- | --- |
| Abstract | 2 | See the PRISMA 2020 for Abstracts checklist |
| Eligibility criteria | 5 | Specify the inclusion and exclusion criteria for the review and how studies were grouped for the syntheses. | page 3; subsection: eligible studies |
| Information sources | 6 | Specify all databases, registers, websites, organisations, reference lists and other sources searched or consulted to identify studies. Specify the date when each source was last searched or consulted. | page 3; subsection: search strategies |
| Search strategy | 7 | Present the full search strategies for all databases, registers and websites, including any filters and limits used. | page 3; subsection: search strategies |
| Selection process | 8 | Specify the methods used to decide whether a study met the inclusion criteria of the review, including how many reviewers screened each record and each report retrieved, whether they worked independently, and if applicable, details of automation tools used in the process. | page 4; subsection: study selection and data extraction |

*(continued)*

|  |  |  |  |
| --- | --- | --- | --- |
| Data collection process | 9 | Specify the methods used to collect data from reports, including how many reviewers collected data from each report, whether they worked independently, any processes for obtaining or confirming data from study investigators, and if applicable, details of automation tools used in the process. | page 4; subsection: study selection and data extraction |
| Data items | 10a | List and define all outcomes for which data were sought. Specify whether all results that were compatible with each outcome domain in each study were sought (e.g. for all measures, time points, analyses), and if not, the methods used to decide which results to collect. | page 4; subsection: study selection and data extraction |
|  | 10b | List and define all other variables for which data were sought (e.g. participant and intervention characteristics, funding sources). Describe any assumptions made about any missing or unclear information. | page 4; subsection: study selection and data extraction |
| Study risk of bias assessment | 11 | Specify the methods used to assess risk of bias in the included studies, including details of the tool(s) used, how many reviewers assessed each study and whether they worked independently, and if applicable, details of automation tools used in the process. | page 4; subsection: data evaluation |
| Effect measures | 12 | Specify for each outcome the effect measure(s) (e.g. risk ratio, mean difference) used in the synthesis or presentation of results. | not applicable: no quantitative synthesis done |
| Synthesis methods | 13a | Describe the processes used to decide which studies were eligible for each synthesis (e.g. tabulating the study intervention characteristics and comparing against the planned groups for each synthesis (item 5)). | not applicable: no quantitative synthesis done |

*(continued)*

|  |  |  |  |
| --- | --- | --- | --- |
|  | 13b | Describe any methods required to prepare the data for presentation or synthesis, such as handling of missing summary statistics, or data conversions. | not applicable: no quantitative synthesis done |
|  | 13c | Describe any methods used to tabulate or visually display results of individual studies and syntheses. | not applicable: no quantitative synthesis done |
|  | 13d | Describe any methods used to synthesize results and provide a rationale for the choice(s). If meta-analysis was performed, describe the model(s), method(s) to identify the presence and extent of statistical heterogeneity, and software package(s) used. | not applicable: no quantitative synthesis done |
|  | 13e | Describe any methods used to explore possible causes of heterogeneity among study results (e.g. subgroup analysis, meta-regression). | not applicable: no quantitative synthesis done |
|  | 13f | Describe any sensitivity analyses conducted to assess robustness of the synthesized results. | not applicable: no quantitative synthesis done |
| Reporting bias assessment | 14 | Describe any methods used to assess risk of bias due to missing results in a synthesis (arising from reporting biases). | not applicable: no quantitative synthesis done |
| Certainty assessment | 15 | Describe any methods used to assess certainty (or confidence) in the body of evidence for an outcome. | not applicable: no quantitative synthesis done |
| **RESULTS** |  |  |  |
| Study selection | 16a | Describe the results of the search and selection process, from the number of records identified in the search to the number of studies included in the review, ideally using a flow diagram. | page 4; last paragraph |
|  | 16b | Cite studies that might appear to meet the inclusion criteria, but which were excluded, and explain why they were excluded. | page 4; last paragraph |
| Study characteristics | 17 | Cite each included study and present its characteristics. | figure 1 |

*(continued)*

|  |  |  |  |
| --- | --- | --- | --- |
| Risk of bias in studies | 18 | Present assessments of risk of bias for each included study. | table 3 |
| Results of individual studies | 19 | For all outcomes, present, for each study: (a) summary statistics for each group (where appropriate) and (b) an effect estimate and its precision (e.g. confidence/credible interval), ideally using structured tables or plots. | table 2 |
| Results of syntheses | 20a | For each synthesis, briefly summarise the characteristics and risk of bias among contributing studies. | page 6; subsection: risk of bias of included studies |
|  | 20b | Present results of all statistical syntheses conducted. If meta-analysis was done, present for each the summary estimate and its precision (e.g. confidence/credible interval) and measures of statistical heterogeneity. If comparing groups, describe the direction of the effect. | not applicable: no statistic syntheses done |
|  | 20c | Present results of all investigations of possible causes of heterogeneity among study results. | not applicable: no statistic syntheses done |
|  | 20d | Present results of all sensitivity analyses conducted to assess the robustness of the synthesized results. | not applicable: no statistic syntheses done |
| Reporting biases | 21 | Present assessments of risk of bias due to missing results (arising from reporting biases) for each synthesis assessed. | page 12; subsection: limitations |
| Certainty of evidence | 22 | Present assessments of certainty (or confidence) in the body of evidence for each outcome assessed. | not applicable for diagnostic test review but for treatment |
| **DISCUSSION** |  |  |  |
| Discussion | 23a | Provide a general interpretation of the results in the context of other evidence. | page 11; discussion 1st paragraph |
|  | 23b | Discuss any limitations of the evidence included in the review. | page 12; subsection: limitations |
|  | 23c | Discuss any limitations of the review processes used. | page 12; subsection: limitations |

*(continued)*

|  |  |  |  |
| --- | --- | --- | --- |
|  | 23d | Discuss implications of the results for practice, policy, and future research. | page 12; 3rd paragraph |
| **OTHER INFORMATION** |  |  |  |
| Registration and protocol | 24a | Provide registration information for the review, including register name and registration number, or state that the review was not registered. | page 3 |
|  | 24b | Indicate where the review protocol can be accessed, or state that a protocol was not prepared. | page 3 |
|  | 24c | Describe and explain any amendments to information provided at registration or in the protocol. | page 12; section: limitations |
| Support | 25 | Describe sources of financial or non-financial support for the review, and the role of the funders or sponsors in the review. | page 13; section: acknowledgements |
| Competing interests | 26 | Declare any competing interests of review authors. | page 13; section: conflicts of interests |
| Availability of data, code and other materials | 27 | Report which of the following are publicly available and where they can be found: template data collection forms; data extracted from included studies; data used for all analyses; analytic code; any other materials used in the review. | page 13; sections: data and materials sharing |

## PRIMSA Abstract Checklist

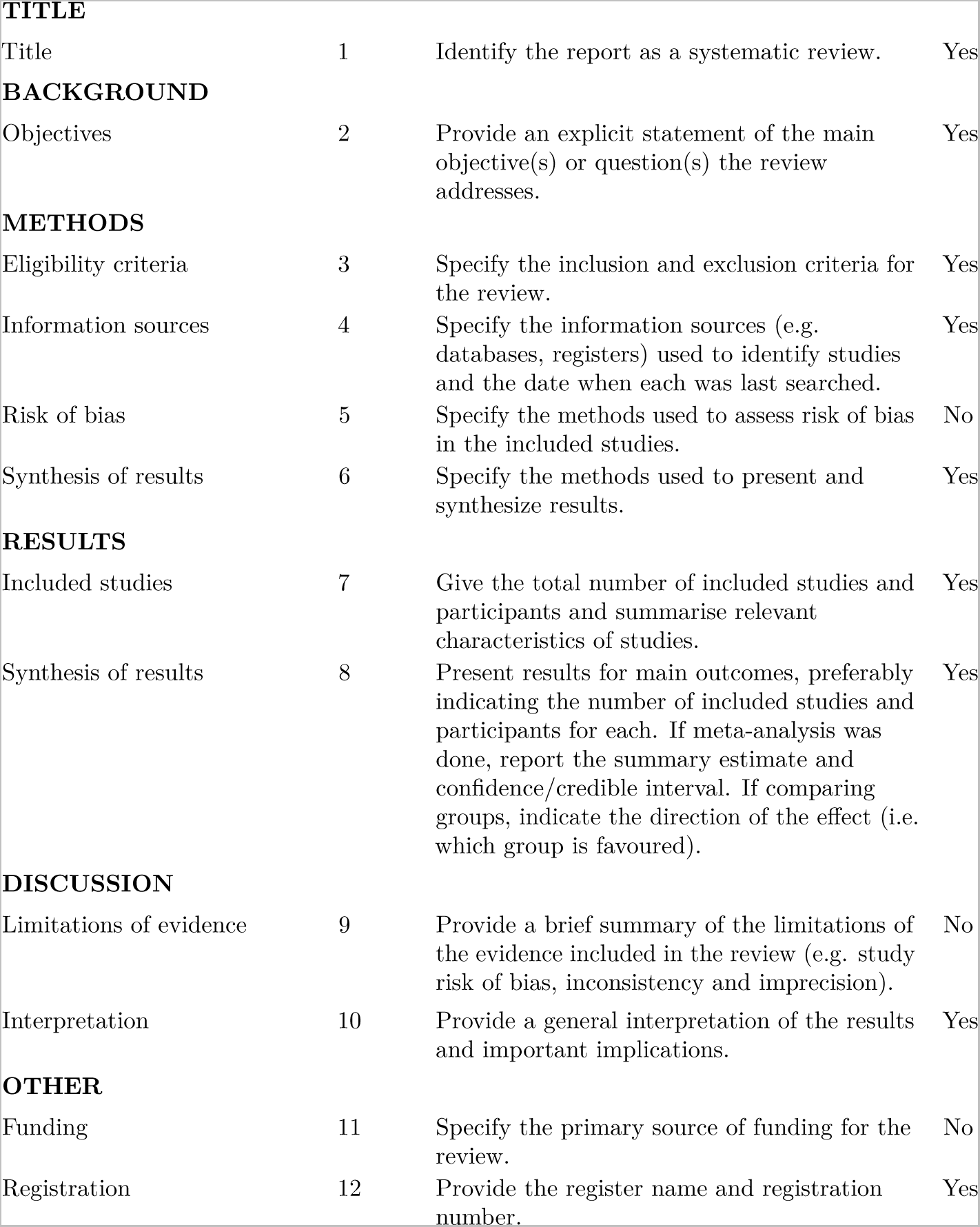

TITLE
|  |  |  |  |
| --- | --- | --- | --- |
| Title | 1 | Identify the report as a systematic review. | Yes |
| BACKGROUND |  |  |  |
| Objectives | 2 | Provide an explicit statement of the main objective(s) or question(s) the review addresses. | Yes |
| Eligibility criteria | 3 | Specify the inclusion and exclusion criteria for the review. | Yes |
| Information sources | 4 | Specify the information sources (e.g. databases, registers) used to identify studies and the date when each was last searched. | Yes |
| Risk of bias | 5 | Specify the methods used to assess risk of bias in the included studies. | No |
| Synthesis of results | 6 | Specify the methods used to present and synthesize results. | Yes |
| Included studies | 7 | Give the total number of included studies and participants and summarise relevant characteristics of studies. | Yes |
| Synthesis of results | 8 | Present results for main outcomes, preferably indicating the number of included studies and participants for each. If meta-analysis was done, report the summary estimate and confidence/credible interval. If comparing groups, indicate the direction of the effect (i.e. which group is favoured). | Yes |
| Limitations of evidence | 9 | Provide a brief summary of the limitations of the evidence included in the review (e.g. study risk of bias, inconsistency and imprecision). | No |
| Interpretation | 10 | Provide a general interpretation of the results and important implications. | Yes |
| OTHER |  |  |  |
| Funding | 11 | Specify the primary source of funding for the review. | No |
| Registration | 12 | Provide the register name and registration number. | Yes |

*From:* Page MJ, McKenzie JE, Bossuyt PM, Boutron I, Hoffmann TC, Mulrow CD, et al. The PRISMA 2020 statement: an updated guideline for reporting systematic reviews. MetaArXiv. 2020, September 14. DOI: 10.31222/osf.io/v7gm2. For more information, visit: www.prisma-statement.org

